# Proteomics Indicates Lactate Dehydrogenase is Prognostic in Acetaminophen-induced Acute Liver Failure Patients and Reveals a Role for LKB1-AMPK Signaling

**DOI:** 10.1101/2021.11.30.21266453

**Authors:** Joel H. Vazquez, Stefanie Kennon-McGill, Stephanie D. Byrum, Samuel G. Mackintosh, Hartmut Jaeschke, D. Keith Williams, William M. Lee, Jonathan A. Dranoff, Mitchell R. McGill, Acute Liver Failure Study Group

**Affiliations:** Dept. of Pharmacology and Toxicology, College of Medicine, University of Arkansas for Medical Sciences, Little Rock, AR 72205; Dept. of Environmental and Occupational Health, Fay W. Boozman College of Public Health, University of Arkansas for Medical Sciences, Little Rock, AR 72205; Dept. of Biochemistry and Molecular Biology, College of Medicine, University of Arkansas for Medical Sciences, Little Rock, AR 72205; Dept. of Pharmacology, Toxicology, and Therapeutics, University of Kansas Medical Center, Kansas City, KS 66160; Dept. of Biostatistics, College of Medicine, University of Arkansas for Medical Sciences, Little Rock, AR 72205; Div. of Digestive and Liver Diseases, Department of Internal Medicine, University of Texas Southwestern Med School, Dallas, TX 75390; Div. of Gastroenterology and Hepatology, Dept. of Internal Medicine, College of Medicine, University of Arkansas for Medical Sciences, Little Rock, AR 72205; Yale University School of Medicine

**Keywords:** Drug-induced liver injury, acute liver injury, liver regeneration, biomarkers

## Abstract

Better biomarkers to predict death early in acute liver failure (ALF) are needed. To that end, we obtained early (study day 1) and later (day 3) serum samples from transplant-free survivors (n=28) and non-survivors (n=30) of acetaminophen (APAP)-induced ALF from the NIH-sponsored Acute Liver Failure Study Group, and from control volunteers (n=10). To identify proteins that increase early in serum during ALF, we selected individuals from this cohort for whom ALT was lower on day 1 than day 3, indicating a time point before the peak of injury (n=10/group). We then performed untargeted proteomics on their day 1 samples. Out of 1,682 quantifiable proteins, 79 were elevated ≥4-fold in ALF patients vs. controls and 23 of those were further elevated ≥4-fold in non-survivors vs. survivors, indicating potential to predict death. Interestingly, the biomarker with best performance was LDH. To confirm the prognostic potential of LDH, we measured activity in all day 1 and 3 samples from all 58 ALF patients. LDH was elevated in the non-survivors vs. survivors on both days. In addition, receiver operating characteristic (ROC) curve analyses revealed that LDH alone performed similarly to the model for end-stage liver disease (MELD), while a combination of MELD and LDH outperformed either alone. Finally, Upstream Analysis of our proteomics data indicated activation of LKB1-AMPK signaling in liver regeneration after APAP overdose and we confirmed that in mice. Overall, we conclude LDH can predict death in APAP-induced ALF and that LKB1-AMPK signaling may be a promising therapeutic target to improve survival.

## INTRODUCTION

Acute liver failure (ALF) is a syndrome of encephalopathy, coagulopathy, and multi-organ dysfunction caused by loss of liver function secondary to acute liver injury. The true incidence of ALF is unknown and estimates vary widely around the world (Hoofnagle *et al*., 1995; Bower *et al*., 2007; Ho *et al*., 2014; Thanapirom *et al*., 2019; Weiler *et al*., 2020), but most agree that it is a relatively rare condition. In the United States, an often-cited figure is 2,000 total cases per year, but crude estimates have ranged from 5.5 to 31.2 cases per million population per year (or around 1,600 to 10,000 cases annually) (Hoofnagle *et al*., 1995; Bower *et al*., 2007). Although ALF is rare, it can be extremely devastating: Despite recent improvements in patient outcomes, overall mortality remains high at around 25-30% (Reuben *et al*., 2016). Currently, the major life-extending treatment for ALF is a liver transplant. However, clinicians must decide quickly when a transplant is needed because the median time from hospital admission or onset of ALF to death is just 5-6 days (Shakil *et al*., 2000; Ostapowicz *et al*., 2002).

Unfortunately, current liver biomarkers help little with the decision to transplant. The primary biomarker of liver injury is serum alanine aminotransferase (ALT). While ALT works well to detect liver injury, it has little to no prognostic value (Christensen *et al*., 1984; Tygstrup and Ranek, 1986; McGill, Staggs, *et al*., 2014; Karvellas *et al*., 2017; Kuroda *et al*., 2021). Although liver function tests like bilirubin, prothrombin time, and the international normalized ratio (INR) correlate better with poor outcomes, they sometimes peak late in ALF progression when a liver transplant is no longer feasible. Finally, prognostic scores like the model for end-stage liver disease (MELD), which includes bilirubin and INR, are helpful but lack sufficient sensitivity and specificity to solely guide patient care (De Clercq *et al*., 2021). Thus, the identification and validation of a new biomarker or panel of biomarkers that can predict death early in ALF, before the peak of injury and dysfunction, could dramatically improve the care of these critically ill patients by allowing early activation of resources necessary for liver transplantation.

Surprisingly, although dozens of papers have reported the discovery and validation of novel, targeted biomarkers for prognosis in ALF (De Clercq *et al*., 2021), untargeted proteomics has never been applied directly to samples from ALF patients. Untargeted methods can measure thousands of markers simultaneously and without bias, so they are powerful tools for biomarker discovery. Here, we used untargeted proteomics analysis to identify biomarker candidates to predict poor outcomes in patients with acetaminophen (APAP)-induced ALF. We then selected one of those biomarkers, lactate dehydrogenase (LDH), for further testing and confirmation. Importantly, LDH is routinely measured in many clinical laboratories and could be immediately implemented as a prognostic biomarker in ALF in routine practice. Finally, we used a reverse translational approach with Ingenuity Pathway Analysis®, Upstream Analysis, and a mouse model of APAP-induced liver injury to explore the mechanistic significance of our untargeted proteomics results.

## METHODS

### ALFSG samples

Serum samples from 28 random transplant-free survivors and 30 random non-survivors of APAP-induced ALF were obtained from the Acute Liver Failure Study Group (ALFSG) biorepository. ALF was diagnosed by ALFSG investigators and defined as INR ≥ 1.5, hepatic encephalopathy, duration of illness <26 weeks, and absence of chronic liver disease. APAP toxicity was determined to be the etiology based on a combination of patient-reported history of APAP overdose, a detectable APAP level documenting ingestion, and aminotransferase level of ≥ 1,000 IU/L. Due to hepatic encephalopathy, consent was obtained from next of kin. Samples were centrifuged at each ALFSG study site to obtain serum and stored at −80°C for later distribution and analysis. Demographic and laboratory data provided with the samples included daily values for serum ALT, AST, total bilirubin (Tbili), prothrombin time (PT), and creatinine (Cre) during hospitalization; age; sex; race; and ethnicity. The internal review board (IRB) at each ALFSG study site (Albert Einstein Medical Center, Baylor University Medical Center, Emory University, Massachusetts General Hospital, Mayo Clinic, Medical University of South Carolina, Mount Sinai School of Medicine, Northwestern University Feinberg School of Medicine, The Ohio State University, Oregon Health Sciences University, University of Alabama – Birmingham, University of Alberta, University of California – San Francisco, University of Kansas Medical Center, University of Michigan Medical Center, University of Nebraska Medical Center, University of Pennsylvania, University of Pittsburgh Medical Center, University of Texas Southwestern Medical Center, University of Washington, Virginia Commonwealth University, Yale University) reviewed and approved the study design and protocol before study initiation and the study was conducted in accordance with the 1975 Declaration of Helsinki.

### Volunteer subjects

Ten volunteers without liver disease and with reported recent therapeutic APAP exposure were recruited at the University of Arkansas for Medical Sciences (UAMS) in Little Rock, AR, USA. Recent therapeutic exposure was considered useful to control for the effects of APAP itself, though not essential for the study. Each subject was informed of the potential risks and benefits of the study and signed a consent form. After enrollment, a blood sample was collected from each subject and serum was separated by centrifugation. The study protocol was reviewed and approved by the UAMS IRB and the study was conducted in accordance with the 1975 Declaration of Helsinki.

### Untargeted Proteomics

Abundant serum proteins were depleted with HighSelect Top14 resin (Thermo) according to the manufacturer’s instructions. Proteins were reduced and alkylated prior to digestion with sequencing grade modified porcine trypsin (Promega) using S-Trap columns (Protifi). Tryptic peptides were then separated by reverse phase XSelect CSH C18 2.5 um resin (Waters) on an in-line 150 × 0.075 mm column using an UltiMate 3000 RSLCnano system (Thermo). Peptides were eluted using a 60 min gradient from 98:2 to 65:35 buffer A:B ratio (buffer A = 0.1% formic acid, 0.5% acetonitrile; buffer B = 0.1% formic acid, 99.9% acetonitrile). Eluted peptides were ionized by electrospray (2.2 kV) followed by mass spectrometric analysis on an Orbitrap Exploris 480 mass spectrometer (Thermo). To assemble a chromatogram library, six gas-phase fractions were acquired on the Orbitrap Exploris with 4 m/z DIA spectra (4 m/z precursor isolation windows at 30,000 resolution, normalized AGC target 100%, maximum inject time 66 ms) using a staggered window pattern from narrow mass ranges using optimized window placements. Precursor spectra were acquired after each DIA duty cycle, spanning the m/z range of the gas-phase fraction (i.e. 496-602 m/z, 60,000 resolution, normalized AGC target 100%, maximum injection time 50 ms). For wide-window acquisitions, the Orbitrap Exploris was configured to acquire a precursor scan (385-1015 m/z, 60,000 resolution, normalized AGC target 100%, maximum injection time 50 ms) followed by 50x 12 m/z DIA spectra (12 m/z precursor isolation windows at 15,000 resolution, normalized AGC target 100%, maximum injection time 33 ms) using a staggered window pattern with optimized window placements. Precursor spectra were acquired after each DIA duty cycle.

### Proteomics Data Analysis

Following data acquisition, data were searched using an empirically corrected library and a quantitative analysis was performed to obtain a comprehensive proteomic profile. Proteins were identified and quantified using EncyclopeDIA (Searle *et al*.) and visualized with Scaffold DIA using 1% false discovery thresholds at both the protein and peptide level. Protein exclusive intensity values were assessed for quality using an in-house ProteiNorm app, a tool for systematic evaluation of normalization methods, imputation of missing values and comparisons of multiple differential abundance methods (Graw *et al*.). Normalization methods evaluated included log2 normalization (Log2), median normalization (Median), mean normalization (Mean), variance stabilizing normalization (VSN) (Huber *et al*.), quantile normalization (Quantile) (Bolstad *et al*.), cyclic loess normalization (Cyclic Loess) (Ritchie *et al*.), global robust linear regression normalization (RLR) (Chawade *et al*.), and global intensity normalization (Global Intensity) (Chawade *et al*.). The individual performance of each method was evaluated by comparing of the following metrics: total intensity, pooled intragroup coefficient of variation (PCV), pooled intragroup median absolute deviation (PMAD), pooled intragroup estimate of variance (PEV), intragroup correlation, sample correlation heatmap (Pearson), and log2-ratio distributions. The normalized data were used to perform statistical analysis using linear models for microarray data (limma) with empirical Bayes (eBayes) smoothing to the standard errors (Ritchie *et al*.). Proteins with an FDR adjusted p-value < 0.05 and a fold change >2 were generally considered significant. For biomarker identification, we focused on biomarkers elevated ≥4-fold in ALF patients vs. controls and further elevated ≥4-fold in non-survivors vs. survivors of ALF.

### Lactate dehydrogenase measurement

Lactate dehydrogenase (LDH) activity was measured using a standard kinetic assay based on the loss of NADH absorbance in the reaction mixture. Briefly, each serum sample was diluted in 1x phosphate-buffered saline (PBS) and mixed with 60 mM potassium phosphate buffer (pH 7.5) containing 0.7 mM pyruvate and 216 mM NADH in at least one well of a 96-well plate. The plate was then placed into a UV/Vis spectrophotometer and absorbance at 340 nm was monitored over 3 minutes in 11 second intervals at 37°C. Activity was calculated using the Beer-Lambert equation and expressed in international units per liter (U/L).

### Pathway analysis

Pathway Analysis and subsequent Upstream Analysis of the untargeted proteomics data were performed using Ingenuity Pathway Analysis® software (Qiagen, Germantown, MD). LogFC cutoffs of -1 to 1 and a p-value cutoff of 0.05 were used in the initial core analysis.

### Animal study

Wild-type male C57Bl/6J mice were acquired from the Jackson Laboratory (Bar Harbor, ME) and housed in a temperature-controlled facility with a 12 h light-dark cycle at the University of Arkansas for Medical Sciences (UAMS). All mice were used in experiments at 8-9 weeks of age. The animals were allowed free access to food and water until the night before APAP administration. Briefly, food was removed overnight beginning at -12 to -16 h, followed by i.p. treatment with 300 mg/kg APAP dissolved in warm 1x phosphate-buffered saline (PBS) at 0 h the next morning, and finally either 20 mg/kg dorsomorphin (Dorso) dissolved in DMSO vehicle or an equal volume of DMSO Veh at 6 h. Food was returned at the time of Dorso/Veh treatment. Blood and liver tissue were collected at 24 h. Alanine aminotransferase (ALT) was measured in serum from the mice using a kit from MedTest Dx (Canton, MI), according to the manufacturer’s instructions. Hematoxylin and eosin (H&E) staining and both immunohistochemistry and immunoblotting for proliferating cell nuclear antigen (PCNA) were performed as previously described (Clemens et al., 2019).

### Statistical analyses

Sensitivity, specificity, likelihood ratios, and post-test probabilities were calculated using standard equations (Deacon, 2009). Data normality was tested using the Shapiro-Wilk test. For normally distributed data, groups were compared using Student’s t-test. For non-normally distributed data, groups were compared using a t-test on ranks. Logistic regression was used to screen for associations between biomarkers and outcome and receiver operating characteristic (ROC) curves were used to visualize the associations. Optimal biomarker cutoffs were determined using logistic regression with sensitivity set at 90%. The equation to combine MELD score and LDH values was MELD-LDH = -1.981 + (0.00008*LDH) + (0.0698*MELD), derived using multiple logistic regression. All statistical analyses were performed in SigmaPlot 12.5 (Systat, San Jose, CA).

## RESULTS

### Participant demographics

Table 1 provides a summary of demographics and clinical laboratory data from the day 1 and day 3 samples for all ALF survivors and non-survivors, as well as data from control subjects.

**Table 1.**
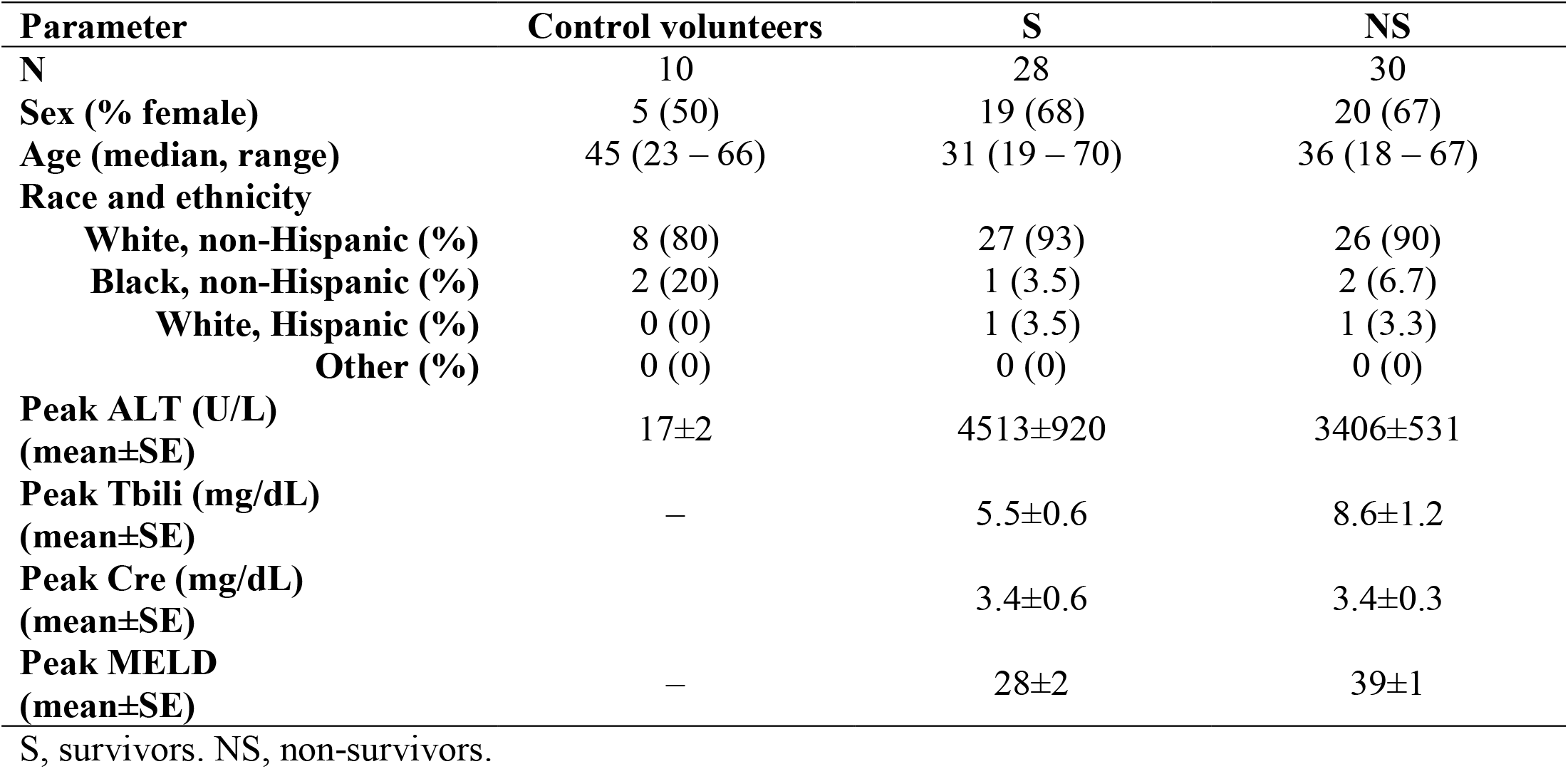
All participant demographics and laboratory data.

| **Parameter** | **Control volunteers** | **S** | **NS** |
| --- | --- | --- | --- |
| **N** | 10 | 28 | 30 |
| **Sex (% female)** | 5 (50) | 19 (68) | 20 (67) |
| **Age (median, range)** | 45 (23 – 66) | 31 (19 – 70) | 36 (18 – 67) |
| **Race and ethnicity** |  |  |  |
| **White, non-Hispanic (%)** | 8 (80) | 27 (93) | 26 (90) |
| **Black, non-Hispanic (%)** | 2 (20) | 1 (3.5) | 2 (6.7) |
| **White, Hispanic (%)** | 0 (0) | 1 (3.5) | 1 (3.3) |
| **Other (%)** | 0 (0) | 0 (0) | 0 (0) |
| **Peak ALT (U/L)**<br>**(mean±SE)** | 17±2 | 4513±920 | 3406±531 |
| **Peak Tbili (mg/dL)**<br>**(mean±SE)** | — | 5.5±0.6 | 8.6±1.2 |
| **Peak Cre (mg/dL)**<br>**(mean±SE)** |  | 3.4±0.6 | 3.4±0.3 |
| **Peak MELD**<br>**(mean±SE)** | — | 28±2 | 39±1 |

S, survivors. NS, non-survivors.

### Discovery proteomics

To identify candidate biomarkers with potential to predict poor outcomes in ALF, we performed untargeted proteomics analysis of the day 1 samples from a subset of 10 participants from each group. Individuals in the subset were chosen because their day 1 samples were collected before ALT peaked during hospitalization, indicating that they were collected early in the progression of ALF, to increase the likelihood that our candidate biomarkers will have utility early in the course of acute liver injury. One-half of each of group consisted of women, and each group included the available black, white, and Hispanic participants to help ensure that the results would be applicable across sex, race, and ethnicity. Table 2 provides demographic and laboratory data for these sub-groups.

**Table 2.** Untargeted proteomics subgroup demographics and laboratory data.

| **Parameter** | **Control volunteers** | **S** | **NS** |
| --- | --- | --- | --- |
| **N** | 10 | 10 | 10 |
| **Sex (% female)** | 5 (50) | 5 (50) | 5 (50) |
| **Age (median, range)** | 45 (23 – 66) | 32 (19 – 46) | 33 (18 – 67) |
| **Race and ethnicity** |  |  |  |
| **White, non-Hispanic (%)** | 8 (80) | 8 (80) | 7 (70) |
| **Black, non-Hispanic (%)** | 2 (20) | 1 (10) | 2 (20) |
| **White, Hispanic (%)** | 0 (0) | 1 (10) | 1 (10) |
| **Other (%)** | 0 (0) | 0 (0) | 0 (0) |
| **Peak ALT (U/L)**<br>**(mean±SE)** | 17±2 | 8275±1494 | 7493±992 |

S, survivors. NS, non-survivors.

Using untargeted proteomics, we were able to quantify 1,682 proteins across all serum samples from the ALF patients and volunteers. To ensure identification of the most robust and promising biomarker candidates, we filtered out proteins that were <4-fold elevated in serum from ALF patients compared to healthy volunteers, leaving 79. We then further filtered out proteins in the list that were <4-fold elevated in samples from ALF non-survivors compared to survivors. This left us with 23 biomarker candidates that were at least 4-fold elevated in the ALF patients overall vs. volunteer controls and again 4-fold higher in ALF survivors vs. non-survivors (**Fig. 1A**). Receiver operating characteristic curve (ROC) analysis further revealed that all 23 candidates had better overall sensitivity and specificity for death than the commonly used model for end-state liver disease (MELD) score in these ALF patients (**Fig. 1B)**.

**Figure 1.**
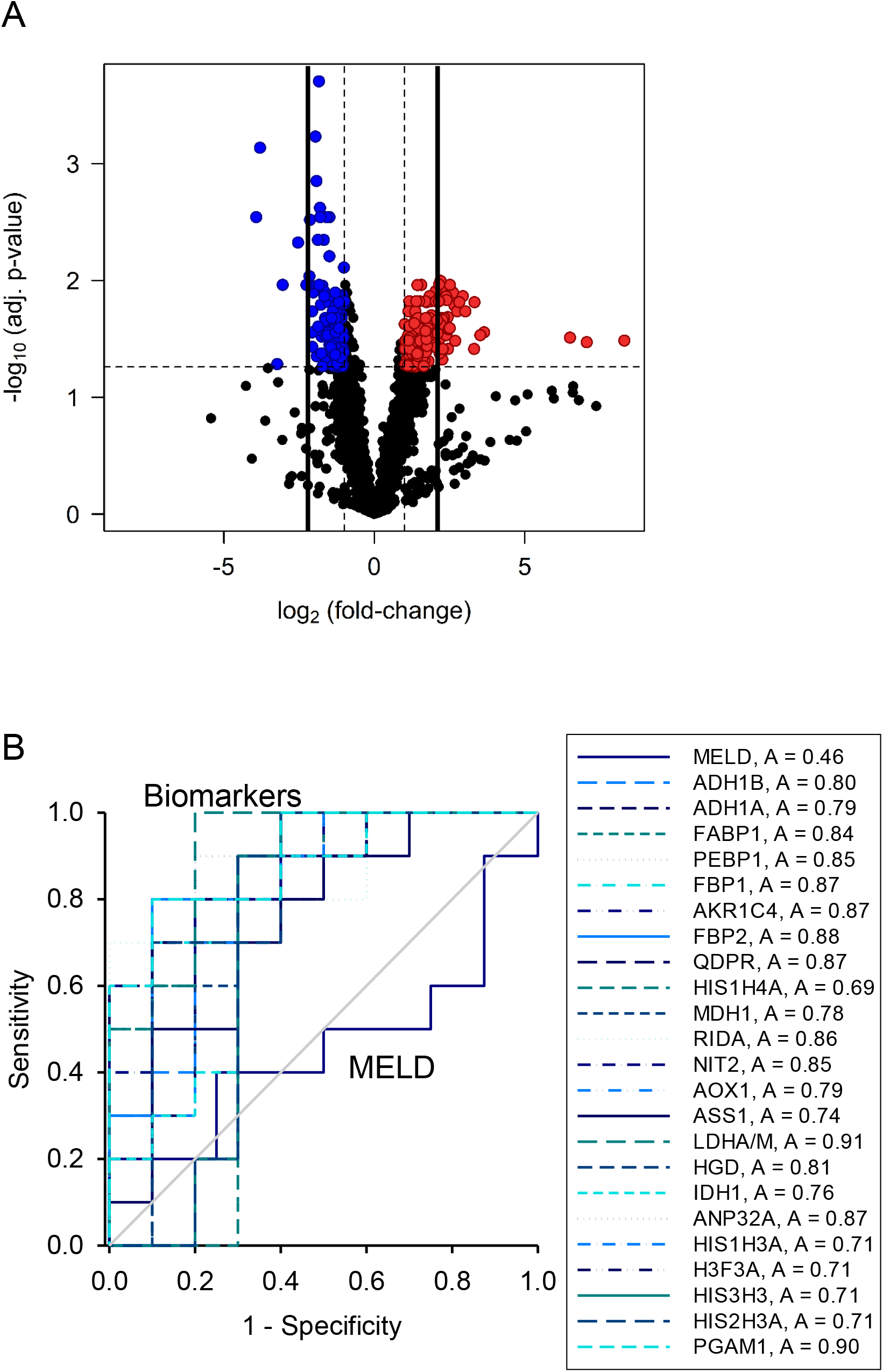
Untargeted proteomics revealed 23 proteins that were ≥4-fold elevated in serum from non-survivors compared to survivors. Day 1 serum samples from survivors (n=10) and non-survivors (n=10) of APAP-induced ALF and healthy controls (n=10) were subjected to untargeted proteomics. (A) Volcano plot displaying results for non-survivors vs. survivors. Each dot represents one serum protein. Numerous proteins were elevated ≥4-fold (right-side solid vertical line) in non-survivors compared to survivors. We focused on 23 that were ≥4-fold elevated in ALF patients overall compared to control subjects for further workup. (B) Receiver operating characteristic (ROC) curves showing sensitivity and specificity for the 23 biomarker candidates at different cutoffs. A: area under the curve.

To determine which of these 23 biomarker candidates are most promising for clinical use, we ranked them based on specificity for death at 90% sensitivity. We prioritized sensitivity and fixed it at 90% based on the rationale that it is better to perform a liver transplant on a patient who does not need one than to deny a new liver to a patient who must have it to survive. Selecting the top candidate from our ranking, then, ensures maximum specificity and positive post-test probability while maintaining high sensitivity for rule-out. **Table 3** shows the rankings for all 23 candidates using these criteria. We were intrigued to discover that LDH-M (encoded by the *LDHA* gene) displayed the greatest potential for clinical use as a prognostic marker to predict death in ALF, as total LDH activity is routinely measured in many clinical laboratories and total LDH released into serum during liver injury should consist almost entirely of LDH-M. Most LDH in cells and serum exists as one of five tetrameric isoforms (LDH1 through LDH5), each composed of various combinations of two major subunits (LDH-M and LDH-H). The dominant tetramer in the liver is LDH-5, which is made up of 4 LDH-M subunits. It follows, therefore, that most LDH released into serum during liver injury is LDH-M.

**Table 3.**
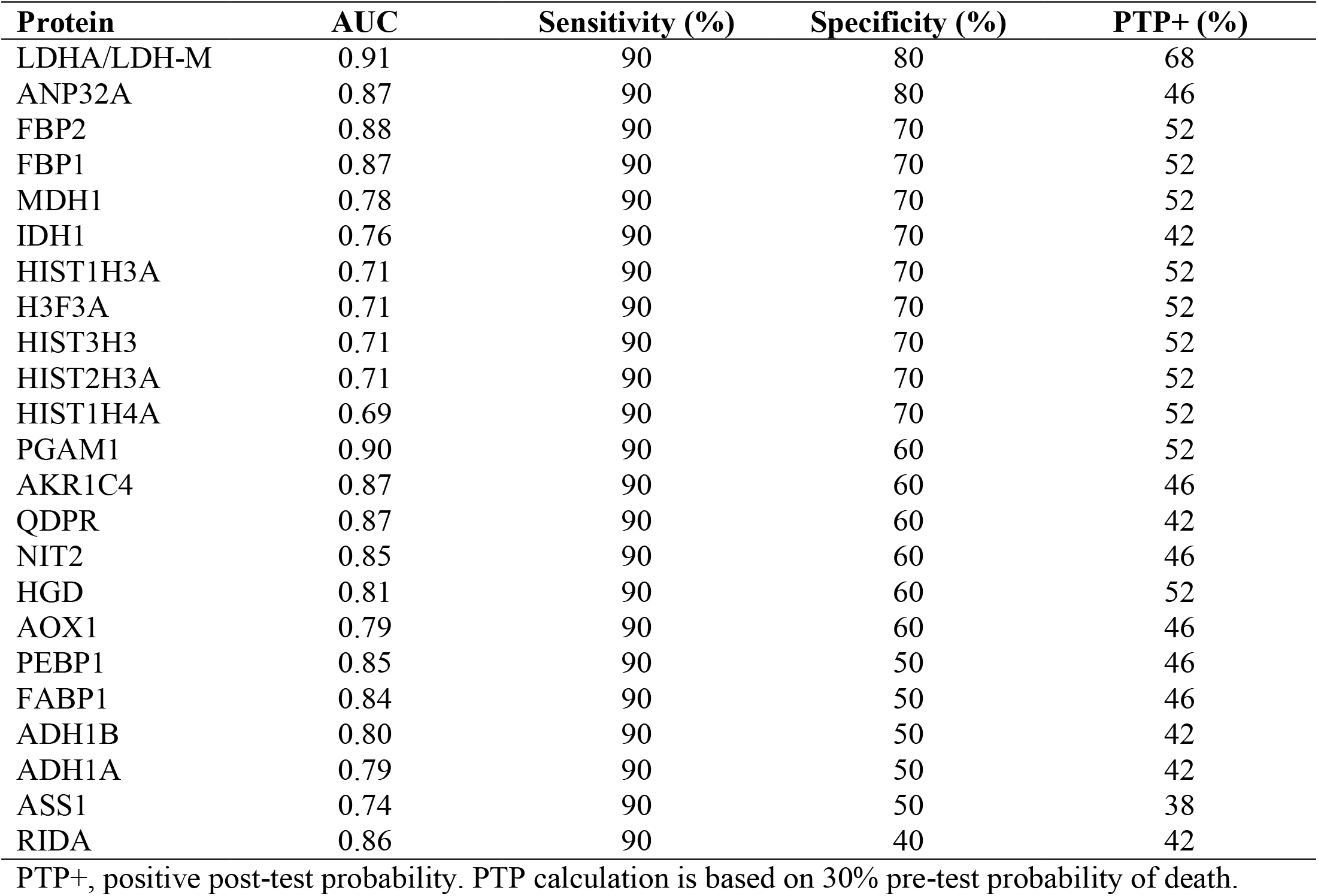
Candidate biomarker ranking.

### Confirmation of lactate dehydrogenase

To further explore the prognostic utility of LDH, we measured total LDH activity in both day 1 and day 3 serum samples from all 60 of the ALF patients. Consistent with our untargeted proteomics results, LDH was significantly higher in serum samples from the non-survivors compared to the survivors on both days 1 and 3 (**Fig. 2A,B**). In addition, both ROC analysis and ranking by specificity at 90% sensitivity revealed that LDH performed similarly overall to the MELD score in our ALF cohort (**Fig. 2C,D**) (**Table 4**). To determine if the combination of LDH and MELD score can predict outcome better than MELD alone, we performed multiple logistic regression and tested the resulting curve fit as a novel prognostic score. Indeed, the novel MELD-LDH score outperformed both LDH alone and MELD alone in the day 3 samples (**Fig. 3**), though not on day 1. These data indicate that LDH may be a more convenient biomarker for prognosis in ALF than MELD due to similar performance without the need for calculation, and further that the combination of MELD and LDH further improves prediction.

**Figure 2.**
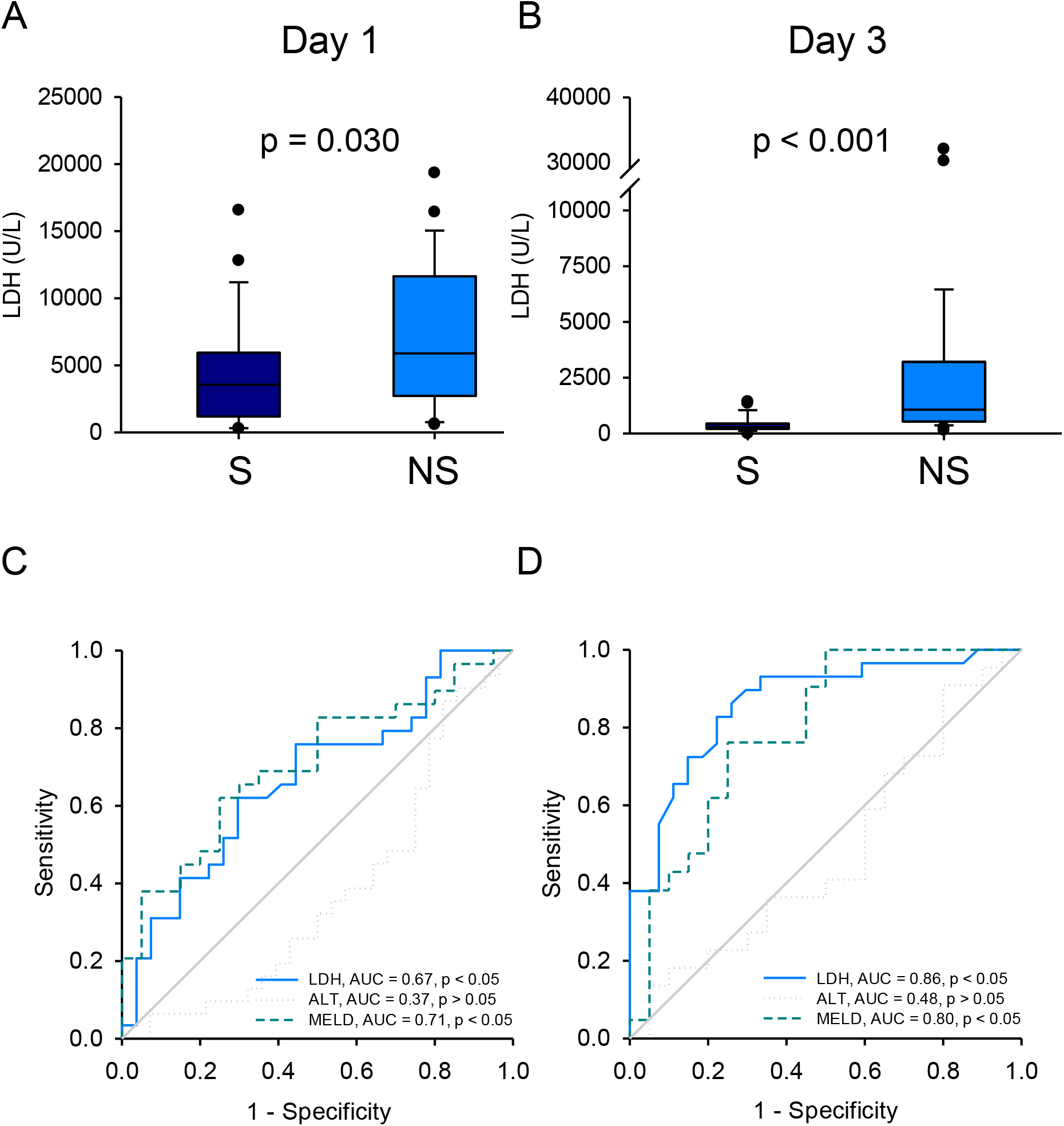
Serum LDH activity was greater in the non-survivors of APAP-induced ALF compared to survivors. Total LDH activity and ALT were measured in serum from all non-survivors (n = 30) and survivors (n = 28) on study days 1 and 3. MELD scores were also obtained when available. (A) LDH activity on day 1. (B) LDH activity on day 3. (C) Receiver operating characteristic (ROC) curves for LDH, ALT, and MELD score on day 1. (D) Receiver operating characteristic (ROC) curves for LDH, ALT, and MELD score on day 3. Differences in AUC values for MELD-LDH vs. MELD and MELD-LDH vs. LDH were statistically significant on day 3. Boxes show the 25^th^ to 75^th^ percentiles. Whiskers show the 10^th^ and 90^th^ percentile values. Lines show median values. Dots represent outliers.

**Table 4.**
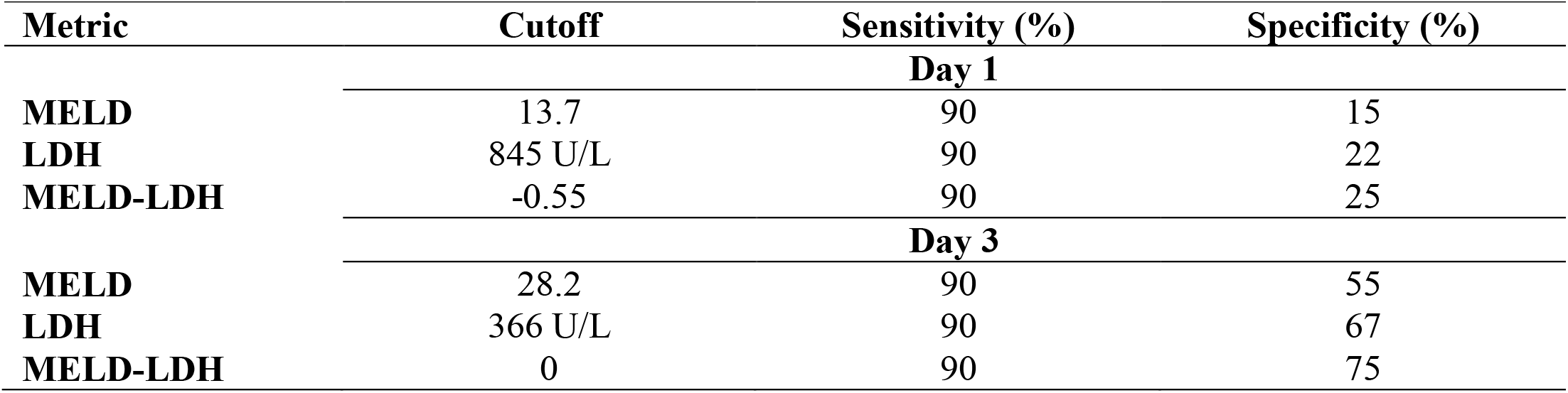
Comparison of MELD-LDH with other metrics.

| **Metric** | **Cutoff** | **Sensitivity (%)** | **Specificity (%)** |
| --- | --- | --- | --- |
|  |  | **Day 1** |  |
| **MELD** | 13.7 | 90 | 15 |
| **LDH** | 845 U/L | 90 | 22 |
| **MELD-LDH** | -0.55 | 90 | 25 |
|  |  | **Day 3** |  |
| **MELD** | 28.2 | 90 | 55 |
| **LDH** | 366 U/L | 90 | 67 |
| **MELD-LDH** | 0 | 90 | 75 |

**Figure 3.**
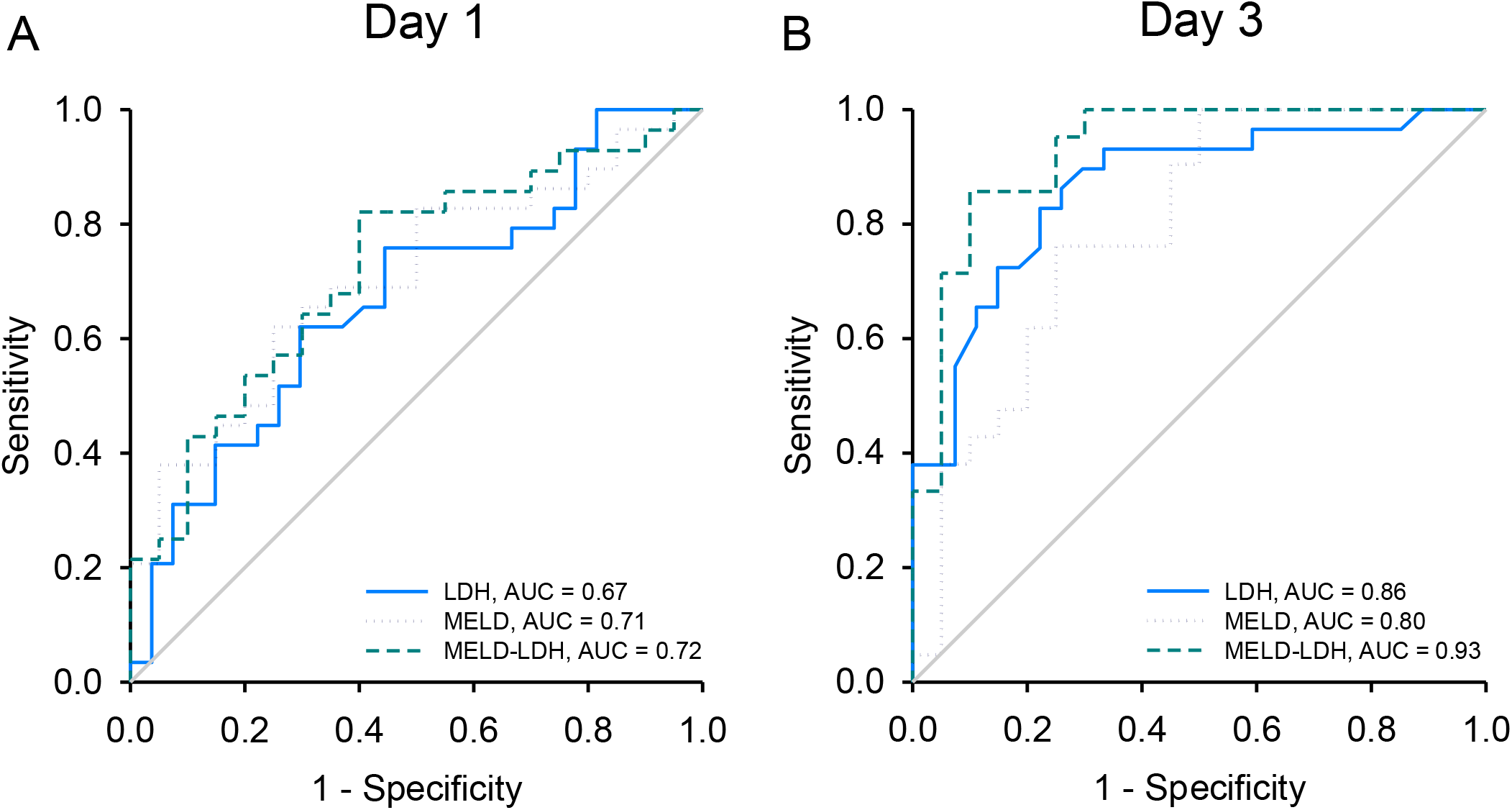
Serum LDH activity improved prediction of death in APAP-induced ALF. Total LDH activity and ALT were measured in serum from all non-survivors and survivors on study days 1 and 3. MELD scores were calculated when possible (n = 20 for survivors, 29 for non-survivors). (A) Receiver operating characteristic (ROC) curves showing sensitivity and specificity for LDH, MELD, and MELD-LDH on day 1. (B) Receiver operating characteristic (ROC) curves showing sensitivity and specificity for LDH, MELD, and MELD-LDH on day 3.

### Pathway analysis

To explore the mechanistic significance of our proteomics data, we next used Ingenuity Pathway Analysis® (IPA) software to perform Upstream Analysis of our untargeted proteomics data. Upstream Analysis is a bioinformatics approach that looks at differences in gene expression or, in this case, protein levels between groups and identifies the likely signaling pathways that were activated to cause those changes in expression based on prior literature. The validity of comparing cell signaling between the survivors and non-survivors using serum is based on two assumptions. First, we assume that hepatocellular damage results in release of most cell proteins into the extracellular milieu such that the serum proteins provide a glimpse of internal cell processes at the time of release. Second, we assume that the extent of cell damage and protein release is similar between the two groups based on the fact that there was no significant difference in serum ALT between the survivors and non-survivors.

Upstream Analysis revealed likely activation of 16 signaling pathways and likely deactivation or suppression of eight others (**Table 5**). Interestingly, several of the pathways that appeared to be activated are known to be involved in cell proliferation and/or liver regeneration. For example, liver kinase b1 (LKB1) signaling promotes hepatocyte proliferation partly through 5’-AMP-activated protein kinase (AMPK) signaling in isolated mouse hepatocytes (Vázquez-Chantada *et al*., 2009). Furthermore, AMPK inhibition blocks liver regeneration after partial hepatectomy in mice (Merlen *et al*., 2014), while chronic activation or over-activation of the aryl hydrocarbon receptor (AhR; another protein in our list) reduces it (Mitchell *et al*., 2006).

**Table 5.**
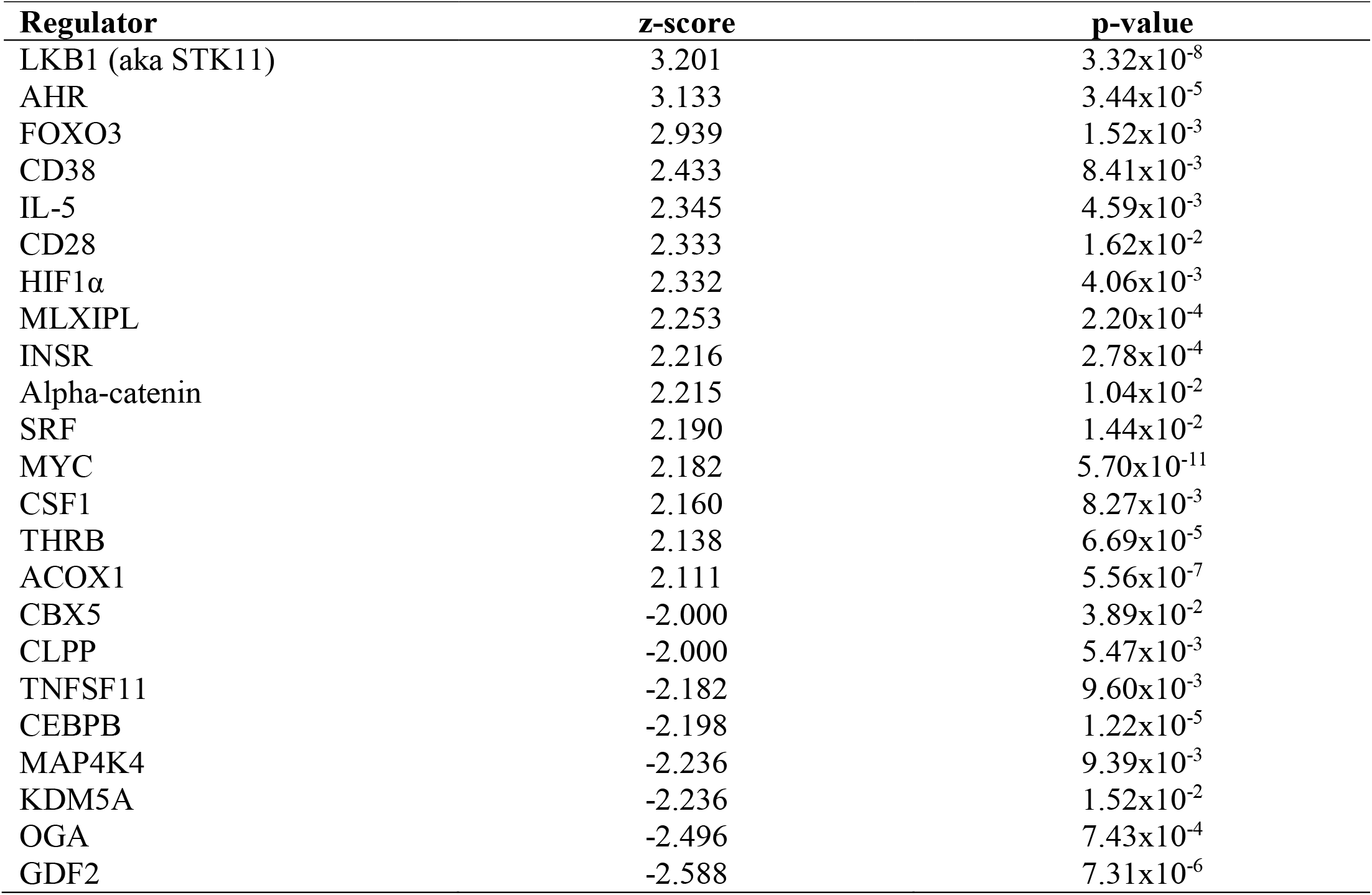
Altered signaling pathways in non-survivors compared to survivors.

Because LKB1 displayed the highest z-score in Upstream Analysis, we wondered if activation of LKB1 signaling in the non-survivors in this study may reflect a last-ditch effort by the liver to repair and regenerate. Accordingly, we hypothesized that inhibition of LKB1-AMPK signaling in mice would reduce or delay liver regeneration. Although direct LKB1 inhibitors are not currently available, there are well-characterized AMPK inhibitors. In addition, it is well-established that LKB1 is the major activator of AMPK (Woods *et al*., 2003; Shaw *et al*., 2004). Therefore, to test our hypothesis, we treated mice with an overdose of APAP at 0 h followed by 20 mg/kg of the AMPK inhibitor dorsomorphin (Dorso) or control vehicle (Veh) at 6 h. The 6 h treatment time point was chosen to avoid any off-target effects on the early APAP metabolism and early liver injury phase thereby focusing the results on regeneration. We then monitored the animals until 24 h, at which time we collected blood and liver tissue from the survivors. The 24 h time point was chosen based on increased mortality in APAP+Dorso-treated mice at this time point in a pilot experiment. Liver injury, as indicated by serum ALT and histology, did not differ between the two groups at 24 h (**Fig. 4A,B**). However, consistent with our hypothesis, AMPK inhibition decreased the marker of liver regeneration proliferating cell nuclear antigen (PCNA) in liver tissue (**Fig. 4B,C**), increased mortality (**Table 6**), and increased symptoms consistent with encephalopathy-like neurological damage (**Table 6**). In addition, Dorso decreased phosphorylation of Raptor, a major target of AMPK (**Fig. 4 C,D**), consistent with inhibition of the AMPK axis. Together, these data are consistent with our hypothesis from the Upstream Analysis results that LKB1-AMPK signaling plays a role in liver regeneration after APAP hepatotoxicity. They also indicate that other pathways known to have a role in cell proliferation that were affected in the non-survivors in our ALF cohort may be important for liver regeneration and survival and should be further studied. We intend to investigate these hypotheses in detail in future studies.

**Figure 4.**
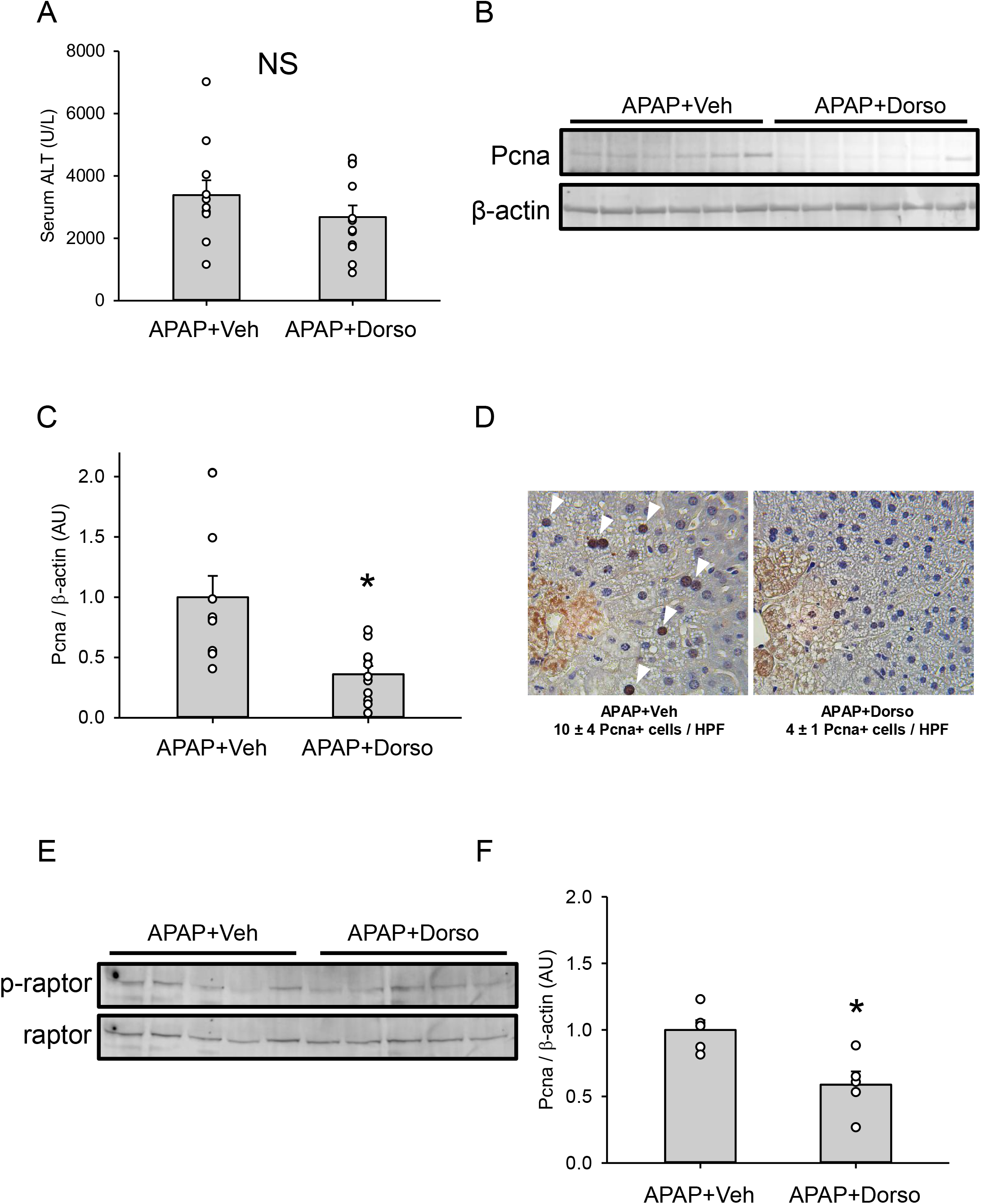
Late post-treatment with dorsomorphin reduced survival and liver regeneration after APAP overdose in mice. Mice were treated with 300 mg/kg APAP at 0 h followed by 20 mg/kg either dorsomorphin (Dorso) or vehicle control (Veh) at 6 h. Blood and liver tissue were collected at 24 h. (A) Serum ALT. (B) Representative immunoblot for proliferating cell nuclear antigen (PCNA) in liver tissue. (C) Densitometry for PCNA immunoblots. (D) Immunohistochemistry for PCNA in liver tissue with counts (positive cells per 400x high-power field [HPF]). White arrowheads point to PCNA-positive nuclei (dark brown, punctate staining pattern). (E) Immunoblot for phosphorylated and total Raptor. (F) Densitometry for Raptor immunoblots. Data are expressed as mean±SE for n = 11-12 total. Note that some individual data points are obscured by overlap. *p<0.05 vs. APAP+Veh.

**Table 6.** ALF endpoints in mouse study with the AMPK inhibitor Dorso.

| **Group** | **Death** | **Neurological symptoms\*** |
| --- | --- | --- |
| **APAP+Vehicle (n = 11)** | 0% | 18% |
| **APAP+Dorsomorphin (n = 12)** | 17% | 92% |

\*Tremor and/or inability to right self when laid on side.

## DISCUSSION

There is a critical need for better tools to predict death and therefore transplant-need early after development of ALF. This is particularly so for APAP-related injury since it evolves very rapidly. A recent study has pointed out that most patients with APAP toxicity have either died, been transplanted, or recovered by day 5 after hospital admission (Reddy *et al*., 2016). Prognostic scores and indexes such as the MELD score and the King’s College Criteria can be helpful, but lack either sensitivity, specificity, or both. MELD score, for example, has limited sensitivity and specificity of around 60-70% each for prediction of death in ALF (De Clercq *et al*., 2021). The King’s College Criteria have better specificity, but similarly poor sensitivity (De Clercq *et al*., 2021), which means that many people who need a transplant to survive would not receive one if relying on the KCC alone to make that choice. Newer scores have been developed with modestly better performance than MELD or KCC, but most rely on uncommon laboratory tests. In addition, scores and indexes can sometimes be onerous for the clinician because they require consideration of multiple laboratory test results and frequently also require manual calculation, as most clinical laboratories do not validate such scores nor provide them in-house. Identification of a single, widely available test that can predict death at least as well as current prognostic scores could facilitate faster life-saving decisions. In the present study, we found that LDH, which is routinely measured in clinical laboratories throughout the world, predicts death as well as the MELD score in APAP-induced ALF. Furthermore, our data indicate that the combination of MELD and LDH to form the novel MELD-LDH score proposed herein improves prediction over either MELD or LDH alone. These results should now be verified in a much larger cohort of patients, which should include ALF of more diverse etiologies.

A point of technical innovation in our work is that we applied untargeted proteomics directly to human samples. Surprisingly, although there has been considerable interest in identifying and developing novel prognostic biomarkers in ALF over the last 20 years (Schmidt and Dalhoff, 2005; McGill, Staggs, *et al*., 2014; Woolbright *et al*., 2014; Karvellas *et al*., 2017; Church *et al*., 2019; Tavabie *et al*., 2021; De Clercq *et al*., 2021), untargeted proteomics has never before been applied directly to patient samples for this purpose. One prior study did use an untargeted proteomics approach in a porcine model of ALF to identify novel ALF biomarkers, but only one of those markers was validated in humans (Wang *et al*., 2017). Interestingly, the latter biomarker was fructose-1,6-bisphosphatase 1 (FBP1), which was also elevated in non-survivors in our cohort. Other potential prognostic biomarkers of interest from our proteomics data included malate dehydrogenase 1 (MDH1), argininosuccinate synthetase 1 (ASS1), and fatty acid-binding protein 1 (FABP1). All three have been proposed as sensitive biomarkers to detect drug-induced liver injury (Schomaker *et al*., 2013; McGill, Cao, *et al*., 2014; Vazquez *et al*., 2020), while there is evidence that FABP1 can also predict death in APAP-induced ALF (Karvellas *et al*., 2017) and poor outcomes other than death in non-APAP ALF (Karvellas *et al*., 2021). Altogether, our proteomics data confirm these earlier results, and indicate that further exploration of FBP1 and FABP1 as prognostic biomarkers is warranted.

In addition to the obvious biomarker applications, our data indicate an important role for LKB1 and LKB1-AMPK signaling in liver regeneration. Although LKB1 is generally thought of as a tumor suppressor that limits cell proliferation, prior studies have yielded conflicting results with regard to the roles of LKB1 and AMPK in liver regeneration. On one hand, it was reported that hepatocyte growth factor (HGF) treatment induces proliferation and phosphorylation of both LKB1 and AMPK in primary mouse hepatocytes and that knockdown of LKB1 reduces the HGF-induced AMPK phosphorylation and proliferation (Vázquez-Chantada *et al*., 2009). Those data strongly indicate that the LKB1-AMPK axis is important for hepatocyte proliferation in mice. Consistent with that, Merlen et al. later reported that AMPKα1 KO mice have delayed liver regeneration after partial hepatectomy (Merlen *et al*., 2014). On the other hand, the latter group also reported that LKB1 KO mice actually have accelerated regeneration in the hepatectomy model (Maillet *et al*., 2018), indicating that LKB1 does indeed function as a suppressor of hepatocyte proliferation in contrast to other data. Here, Upstream Analysis indicated greater activation of LKB1 in non-survivors compared to survivors. If it is correct that the kinase promotes proliferation, then this could be a sign that the hepatocytes are trying to recover but failing. However, if it is correct that the kinase suppresses regeneration, then it could actually be the cause of the patient mortality. To explore this further, we treated mice with the AMPK inhibitor Dorso late after APAP overdose and measured survival and liver regeneration. Our observation that Dorso reduced survival and PCNA expression is consistent with the former hypothesis that LKB1-AMPK signaling promotes regeneration, at least in the case of APAP hepatotoxicity. In addition to mediating HGF signaling, activation of AMPK could be beneficial by promoting autophagy. The Raptor-containing mechanistic target of rapamycin complex 1 (mTORC1) inhibits autophagy and AMPK prevents that by phosphorylating Raptor (Herzig and Shaw, 2018). Indeed, we observed reduced Raptor phosphorylation with Dorso treatment. In addition, AMPK can phosphorylate autophagy mediators like ULK1, which activates them (Herzig and Shaw, 2018). Consistent with this hypothesis, recent evidence demonstrates that blocking mTOR improves liver regeneration and survival in APAP hepatotoxicity in mice (Sun *et al*., 2021). In any case, these data indicate that promoting AMPK activation could be a promising therapeutic approach to improve transplant-free survival in ALF.

Although our results are interesting, this study does suffer from a few weaknesses. First, our ALF groups were relatively small. Additional studies with larger cohorts are needed. Second, our ALF groups included only those with APAP-induced ALF. It is not yet clear how the results would translate to non-APAP ALF. And third, our investigation of the role of LKB1-AMPK signaling in liver regeneration in mice was intended to be preliminary, as the primary focus of this paper is on biomarkers. For example, we were forced to rely on an AMPK inhibitor to test our hypothesis. Although Dorso is a well-characterized and specific AMPK inhibitor, it is possible that LKB1 has other signaling roles during liver regeneration that would offset the effects mediated by AMPK and alter our mechanistic conclusions. Studies with KO mice that include greater mechanistic detail are still needed.

## CONCLUSIONS

Overall, we conclude that LDH is a promising, widely-available biomarker to predict poor outcome in APAP-induced ALF. Further, in our cohort of ALF patients, the combination of MELD and LDH predicted outcome as well as MELD score alone on day 1 and modestly better on day 3. In addition, LKB1-AMPK signaling is likely critical for liver regeneration in these patients based on our Upstream Analysis and mouse data, such that promoting AMPK activity may be a novel approach to improve survival in late-presenting APAP overdose patients. In future human studies, we will verify our results using larger patient cohorts with a variety of ALF etiologies, while in future rodent studies we will explore the role of LKB1 signaling in detail using more rigorous approaches. Lastly, the MELD-LDH score should undergo rigorous testing to assess survival in separate ALF populations because it may have the ability to improve patient outcomes in one of the most difficult clinical conditions faced by hepatologists.

## Data Availability

All data produced in the present study are available upon reasonable request by academic researchers to the authors.

